# Measuring COVID-19 and Influenza in the Real World via Person-Generated Health Data

**DOI:** 10.1101/2020.05.28.20115964

**Authors:** Nikki Marinsek, Allison Shapiro, Ieuan Clay, Ben Bradshaw, Ernesto Ramirez, Jae Min, Andrew Trister, Yuedong Wang, Tim Althoff, Luca Foschini

## Abstract

**Background:** Since the beginning of the COVID-19 pandemic, data from smartphones and connected sensors have been used to better understand presentation and management outside the clinic walls. However, reports on the validity of such data are still sparse, especially when it comes to symptom progression and relevance of wearable sensors.

**Objective:** To understand the relevance of Person-Generated Health Data (PGHD) as a means for early detection, monitoring, and management of COVID-19 in everyday life. This type of data include quantifying prevalence and progression of symptoms from self-reports as well as changes in activity and physiological parameters continuously measured from wearable sensors, and contextualizing findings for COVID-19 patients with those from cohorts of flu patients.

**Design, Setting, and Participants:** Retrospective digital cohort study of individuals with a self-reported positive SARS-CoV-2 or influenza test followed over the period 2019-12-02 to 2020-04-27. Three cohorts were derived: Patients who self-reported being diagnosed with flu prior to the SARS-CoV-2 pandemic (N=6270, of which 1226 also contributed sensor PGHD); Patients who reported being diagnosed with flu during the SARS-CoV-2 pandemic (N=426, of which 85 also shared sensor PGHD); and patients who reported being diagnosed with COVID-19 (N=230, of which sensor PGHD was available for 41). The cohorts were derived from a large-scale digital participatory surveillance study designed to track Influenza-like Illness (ILI) incidence and burden over time.

**Exposures:** Self-reported demographic data, comorbidities, and symptoms experienced during a diagnosed ILI episode, including SARS-CoV-2. Physiological and behavioral parameters measured daily from commercial wearable sensors, including Resting Heart Rate (RHR), total step count, and nightly sleep hours.

**Main Outcomes and Measures:** We investigated the percentage of individuals experiencing symptoms of a given type (e.g. shortness of breath) across demographic groups and over time. We examined illness duration, and care seeking behavior, and how RHR, step count, and nightly sleep hours deviated from expected behavior on healthy days over the course of the infection episode.

**Results:** Self-reported symptoms of COVID-19 present differently from flu. COVID-19 cases tended to last longer than flu (median of 12 vs. 9 days), are uniquely characterized by chest pain/pressure, shortness of breath, and anosmia. The fraction of elevated RHR measurements collected daily from commercial wearable devices rise significantly in the 2 days surrounding ILI symptoms onset, but does not appear to do so in a way specific to COVID-19. Steps lost due to COVID-19 persists for longer than for flu.

**Conclusion and Relevance:** PGHD can be a valid source of longitudinal real world data to detect and monitor COVID-19-related symptoms and behaviors at population scale. PGHD may provide continuous, near real-time feedback to intervention effectiveness that otherwise requires waiting for symptoms to develop into contacts with the healthcare system. It has also the potential to increase pre-test probability of other downstream diagnostics. To effectively leverage PGHD for participatory surveillance it is crucial to invest in the creation of trusted, long-term communication channels with individuals through which data can be efficiently collected, consented, and contextualized, while protecting the privacy of individuals and ultimately facilitating the transition in and out of care.

## Introduction

The emergence of the novel SARS-CoV-2 (COVID-19) and subsequent rapidly expanding pandemic has created significant gaps in our understanding of the prevalence of symptoms among individuals with COVID-19.

Multi-disciplinary teams of physicians, data scientists, clinical informaticians, epidemiologists and many others around the world have engaged in using real-world data collected at point of care to help answer key questions around management of COVID-19 patients (see (1) for a US-centric overview of initiatives and open questions). At the same time, data from individuals in the context of participatory syndromic surveillance for COVID-19 (2–4) are being collected via smartphone apps around the world to perform hotspot detection and show promise in understanding symptom presentation and prevalence outside the clinic walls (5). In addition to self-report, recent literature suggests that data from commercial sensors may be used for large scale surveillance of influenza outbreak, based on the fact that physiologic measures provided by the sensors (e.g., RHR) (6–8) and temperature (9) change in the presence of an infection. Several efforts are currently underway exploring the potential of wearable technology in support of syndromic surveillance aimed at the COVID-19 pandemic (10–12), however, no data yet exists on quantifying the link between COVID-19 disease and changes in physiological and behavioral parameters over the course of the disease. While specific clinical symptoms such as shortness of breath, fever, and dry cough have become hallmark characteristics of the disease (13), symptoms presentation and patient behavior outside the clinic walls has received little attention. Lack of testing means we cannot build a canonical symptom presentation, which significantly undermines our ability to track, predict and control disease progression and manage critical care.

Using a large-scale digital participatory surveillance study designed for the purpose of monitoring ILI over the 2019–2020 influenza season, we present data collected from a cohort of individuals who have self-reported being diagnosed by a medical provider with flu or COVID-19. A subset of this cohort provided daily physiological and behavioral data derived via wearable activity monitors (daily RHR, daily step count, and nightly sleep hours) allowing for the explicit linkage of illness onset and continuously measured physiological and behavioral parameters. Our contributions are twofold. First, we present data on the progression of COVID-19 symptoms in everyday life and contextualize those by comparing them with seasonal influenza. Second, we show that changes in physiological signals such as RHR are associated with the onset of symptoms, though they may not be specific to the type of ILI. To our knowledge, this is the first study that looks at longitudinal symptom reports of COVID-19 patients. It is also the first study presenting symptom reports linked to wearable data at the individual level for ILI (flu or COVID-19), enabling temporal alignment of symptom reports with correspondent changes in wearable data.

## Dataset Description

Evidation Health currently supports a mobile consumer application called Achievement (14) that rewards members based on completing health-related behaviors and participating in research by sharing Person-Generated Health Data (PGHD). Achievement members can connect activity trackers and health applications and authorize Achievement to automatically and continuously ingest connected data streams (15). Achievement has an active user base of 3.7M individuals that are economically, demographically, and geographically diverse, and enables rapid (16) recruitment of participants.

Since 2017, Achievement has been used to run a participatory ILI surveillance program, examining annual waves of Influenza virus infections (17). The 2019–2020 version of the program consists of sending a weekly one-click survey to all Achievement members that asks if the individual experienced any flu-like symptoms in the past 7 days. Individuals who answer “yes” are immediately sent to a questionnaire, which contains questions about the dates of illness onset and/or recovery, symptom experiences in the previous 7 days, healthcare interactions and outcomes, medications, and household characteristics. On 2020–03–30, the questionnaire was updated to include questions that specifically address COVID-19, including questions about COVID-19 diagnosis, testing, and social distancing measures, as well as an expanded list of symptoms (consisting of shortness of breath, chest pain, and anosmia). The contents of the original and updated questionnaires are included in Supplementary Note 2 in the Appendix. Since participants were sent a one-click survey every week, participants could submit multiple survey responses for the same ILI event. Responses to the original and updated surveys, collected between 2019–12–02 and 2020–04–27, comprise the initial survey dataset and include a total of 194,401 responses from 85,558 unique participants.

In addition to agreeing to participate and sharing their survey responses, participants agreed to share their activity data from connected wearable sensors. The sensors data analyzed in this project consisted of minute-by-minute step counts, RHR recordings, and sleep states from 2019–11–01 to 2020–05–13 for the subset of participants with Fitbit sensors connected to the Achievement platform.

## Methods

### Survey Preparation

The pipeline for preparing the surveys for analysis is illustrated in Supplementary Figure S1 (a) and summarized here. Initial survey cleaning consisted of excluding all survey responses with self-reported illness onset dates or recovery dates that occurred 30 or more days before the survey completion date, excluding surveys with invalid illness onset and/or recovery dates (defined as dates occurring after the survey date or responses in which the illness recovery date occurred before the illness onset date), removing multiple survey responses from the same participant on the same day, and de-duplicating identical sets of survey responses. This filtering process reduced the dataset from 194,401 survey responses from 85,558 unique participants to 146,133 responses from 71,556 unique participants.

Since participants could submit multiple survey responses for the same ILI event, distinct ILI events were inferred by merging survey responses from the same participant when the dates encompassing self-reported illness onset through recovery overlapped or were separated by no more than two days. The reconciliation process for merging individual question responses is described in Supplementary Note 3 in the Appendix. After excluding participants with five or more ILI events or multiple diagnosed ILI events (to remove participants with possible erroneous or fraudulent responses), the analysis set was reduced to a subset of 6,926 ILI events with self-reported confirmed diagnosis, each corresponding to a different participant.

The analysis set was supplemented with demographic information (if available) from a different general health survey that included information about gender, age, body mass index (BMI), ethnicity, race, and pre-existing health conditions. Since the general health surveys were completed at different times as the ILI survey, there may be slight discrepancies in time-variant demographic data, such as age, BMI, and health conditions.

### Cohort Definition

All participants included in the analysis self-reported seeking medical care and being diagnosed with flu and/or COVID-19 by a healthcare provider (N=6,926). As shown in Figure 1, these participants were divided into three cohorts. Participants who completed the amended survey and self-reported being diagnosed with COVID-19 were assigned to the COVID-19 cohort (N=230). Participants who completed the updated survey and reported being diagnosed with the flu were assigned to the Non-COVID-19 Flu cohort (N=426). This group allows a comparison of COVID-19 and flu cases that is not confounded by time of year, time since survey completion, survey content, or recent large-scale societal changes, such as shelter-in-place orders or changes within the healthcare system. Participants who reported being diagnosed with both flu and COVID-19 (N=83) were assigned to the COVID-19 cohort, under the rationale that some individuals may consider COVID-19 to be a type of flu, and the relative order in the questionnaire (flu preceding COVID-19). Finally, participants who reported being diagnosed with flu in the original survey were assigned to the Pre-COVID-19 Flu cohort (N=6270). These cases spanned the 2019–2020 flu season before the COVID-19 outbreak, and are included to provide a comparison to a larger group of canonical flu events.

**Fig. 1.**
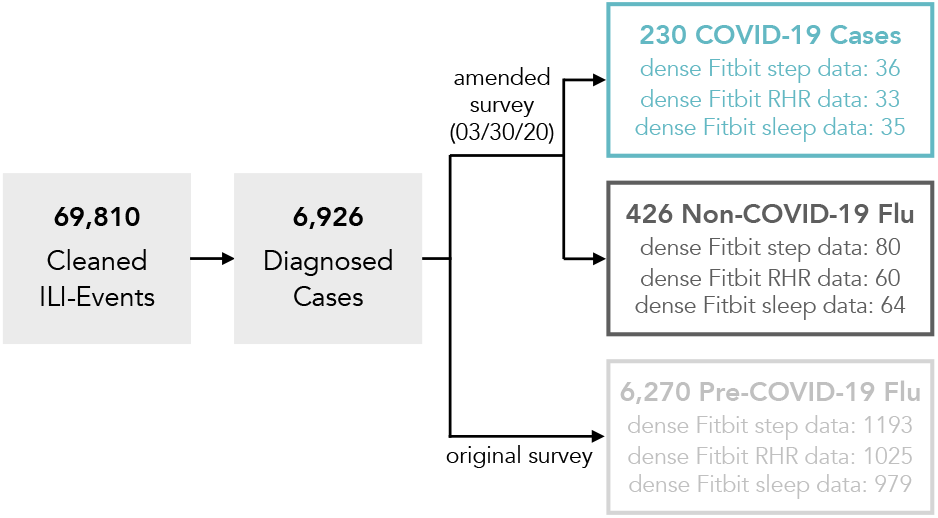
Definition of analysis cohorts. All participants included in the analyses reported that they sought medical care and were diagnosed with either Flu or COVID-19 by a healthcare provider. Participants who indicated they were diagnosed with both Flu and COVID-19 (N=83) were assigned to the COVID-19 cohort.

### Statistical Testing

A two-step statistical testing procedure was used to test for differences in demographics, healthcare care-seeking behavior, medical outcomes, and symptoms among the three cohorts. First, for each sub-analysis (i.e., demographics, medical care-seeking, and symptom prevalence), a series of chi-squared tests of independence were performed to test for an association between the three cohorts and the different possible outcomes for each category. A Bonferroni correction was applied to adjust for running multiple chi-squared tests in each sub-analysis.

Second, follow-up two-proportion z-tests were performed to test for differences in proportions for each outcome and each pair of cohorts. These follow-up tests were only performed for the categories with significant cohort differences as determined by the chi-squared tests.

### Wearable Sensor Data Preparation

Of the 6,926 participants with diagnosed ILI events, 4,778 (69%) have shared as part of the study data from at least one wearable device connected to the Achievement platform: 2,582 (37%) participants had connected Apple Watches, 2,166 (31%) had connected Fitbit devices, 420 (6%) had connected Garmin devices, 123 (2%) had connected Withings devices, and 17 (0.2%) had connected Misfit devices. We focus the analysis of sensor data on the subset of participants with connected Fitbit devices, consisting of minute-by-minute steps, heart rate recordings, and sleep states, were available for a subset of study participants. This data was collected from 2019–11–01 through 2020–05–13 and analyzed to investigate the impact of COVID-19 and flu on everyday behavior and physiology.

Since the sensor data was collected passively in real-world settings, daily sensor wear-time varied across participants and study days. We implemented a three step procedure to enforce adequate data density around each ILI event prior to analysis. First, we estimated if the sensor was worn for each participant for each minute in the study period. Periods of non-wear-time were defined as 180 or more consecutive minutes of zero steps or null RHR recordings. Second, days with 10 or more hours of sensor wear-time were tagged as valid for analysis. For the sleep data, days with a main sleep period recorded by Fitbit were considered valid. Third, the analysis set was restricted to only include participants with 1) at least 10% of valid days for each day of the week in the baseline period (defined as all participant-days that occurred outside the window of 10 days prior to and 20 days after illness onset) and 2) at least 50% valid days in the time period surrounding the ILI event (defined as all days within the window of 10 days prior to 20 days after illness onset). Dense sensor data was available for 41 COVID-19 patients (36 with steps, 33 with RHR, and 35 with sleep), 85 Non-COVID-19 Flu patients (80 with steps, 60 with RHR, and 64 with sleep), and 1226 Pre-COVID-19 Flu patients (1193 with steps, 1025 with RHR, and 979 with sleep). Sensitivity analysis on the valid day thresholds was conducted and results did not change significantly when removing the requirement of having 10% of valid days for each day of week, or lowering the percentage of individual valid days to as low as 30%. The pipeline for preparing the wearable data for analysis is illustrated in Supplementary Figure S1 (b).

### Elevated RHR Prevalence

Similarly to previous work (8), we examined the fraction of each cohort with elevated RHR in the days preceding and following ILI onset. First, days with no RHR recordings were imputed in order to ensure that the cohorts were the same across days of interest. Imputed RHR values were generated from predictions of a mixed effects regression model that was fit to all participant-days that RHR was recorded. The model specified fixed-effects for the week of the year to control for time of year effects (more specifically, this consisted of three terms for the 1st, 2nd, and 3rd expansions of an ordinal variable for week of flu season), a categorical fixed effect for the day of the week to account for differences in activity patterns by day of week, a fixedeffect for the average activity level in the participants’ state of residence to control for different state-wide shelter-in-place and social distancing measures, and a random-intercept for each participant’s baseline activity level to control for individual differences in activity levels. The model was fit to all participant-days with a RHR recording using the lme4 package for R (18). Note that the imputed values were used only to fill days when RHR was not recorded, on all other days the observed value was used.

Next, in order to account for individual differences in RHR when defining thresholds for elevated RHR, RHR values were converted to z-scores using each participant’s RHR mean and standard deviation across all days. The fraction of each cohort with elevated RHR was computed for the days surrounding the ILI event, defined as 10 days prior to 20 days after ILI onset. Elevated RHR was defined as being greater than 1 standard deviation above the participant’s mean RHR. Two-proportion z-tests were performed to answer the following two questions: 1. Does a greater fraction of the COVID-19 cohort have elevated RHR in the days surrounding ILI onset (defined as Days –2 to 2^1^) compared to baseline days prior to ILI onset (defined as Days –10 to –5^2^) and 2. Does the fraction of participants with elevated RHR surrounding ILI onset differ between COVID-19 and Flu cohorts?

### Behavioral and Physiological Changes During ILI Events

In order to characterize daily changes associated with COVID-19 and flu events, we measured deviations from typical healthy measurements (RHR, step count, sleep hours) that occurred while participants were ill. We used a model on symptom-free days (conservatively assumed all days excluding the range 10 days before symptoms onset and within 20 days after symptoms onset) to generate individualized estimates of daily measurements that would have been recorded in the counter-factual scenario that the participant did not fall ill, and then computed the excess, defined as the difference (observed – estimated), on the days surrounding symptoms onset, and finally report the excess as a measure of deviations from expected typical measurements. The symptom-free day model was a mixed effects regression model with the same specification as what was used to impute missing RHR values in the previous analysis. The key difference in this analysis was that, in order to generate estimates based only on assumed symptom-free days, we excluded all data within 10 days before symptoms onset and within 20 days after symptoms onset when fitting the model. In order to visualize the time course of behavioral changes during COVID-19 and flu events, for each cohort we fit generalized additive mixed models with spline smoothing functions and random intercepts to the daily excess time series using the mgcv package for R (20). This procedure was performed three separate times, once for each of the channels considered: daily total step counts, daily RHR, and total daily sleep minutes.

## Results

In the following section, we compare the presentation of diagnosed COVID-19 cases (N=230) to two groups of diagnosed flu cases: Non-COVID-19 Flu cases (N=426), which occurred in the same time frame as the COVID-19 cases, and Pre-COVID-19 Flu (N=6270), which occurred earlier in the 2019–2020 flu season before the outbreak of COVID-19.

### Comparison of Demographic Trends

A demographic summary of the three cohorts is provided in Table 1. A chi-squared test of independence was performed for each demographic category to test for significant differences across the full cohorts. Age-group and race were the only demographics that differed significantly among the cohorts after applying a Bonferroni correction to adjust for performing five comparisons (age group: p =.008, race: p<.001). Compared to the Pre-COVID-19 Flu cohort, the COVID-19 cohort was less likely to be White/Caucasian (63.9% vs. 70.0%) and more likely to be Asian or Pacific Islander (9.6% vs. 4.6%) (follow-up two-proportion z-tests, p =.047 and p<.001, respectively). The COVID-19 cohort was also more likely to prefer not to report race than both the Non-COVID-19 Flu and Pre-COVID-19 Flu cohorts (p=.018 and p<.001, respectively). In follow-up comparisons, the proportion of the COVID-19 cohort belonging to each age group was not significantly different than the Non-COVID-19 Flu nor the Pre-COVID-19 Flu cohorts, however a greater proportion of the Non-COVID-19 Flu cohort was aged 55 or older compared to the Pre-COVID-19 Flu cohort (7.5% vs. 3.8%, two-proportion z-test, p =.001). The demographics of the analyzed cohorts differs from what described in the literature for medically attended ILI events for the general US population (21). The sample should be reweighed (22) before meaningful comparisons can be made with a target population with different demographics characteristics.

**Table 1.**
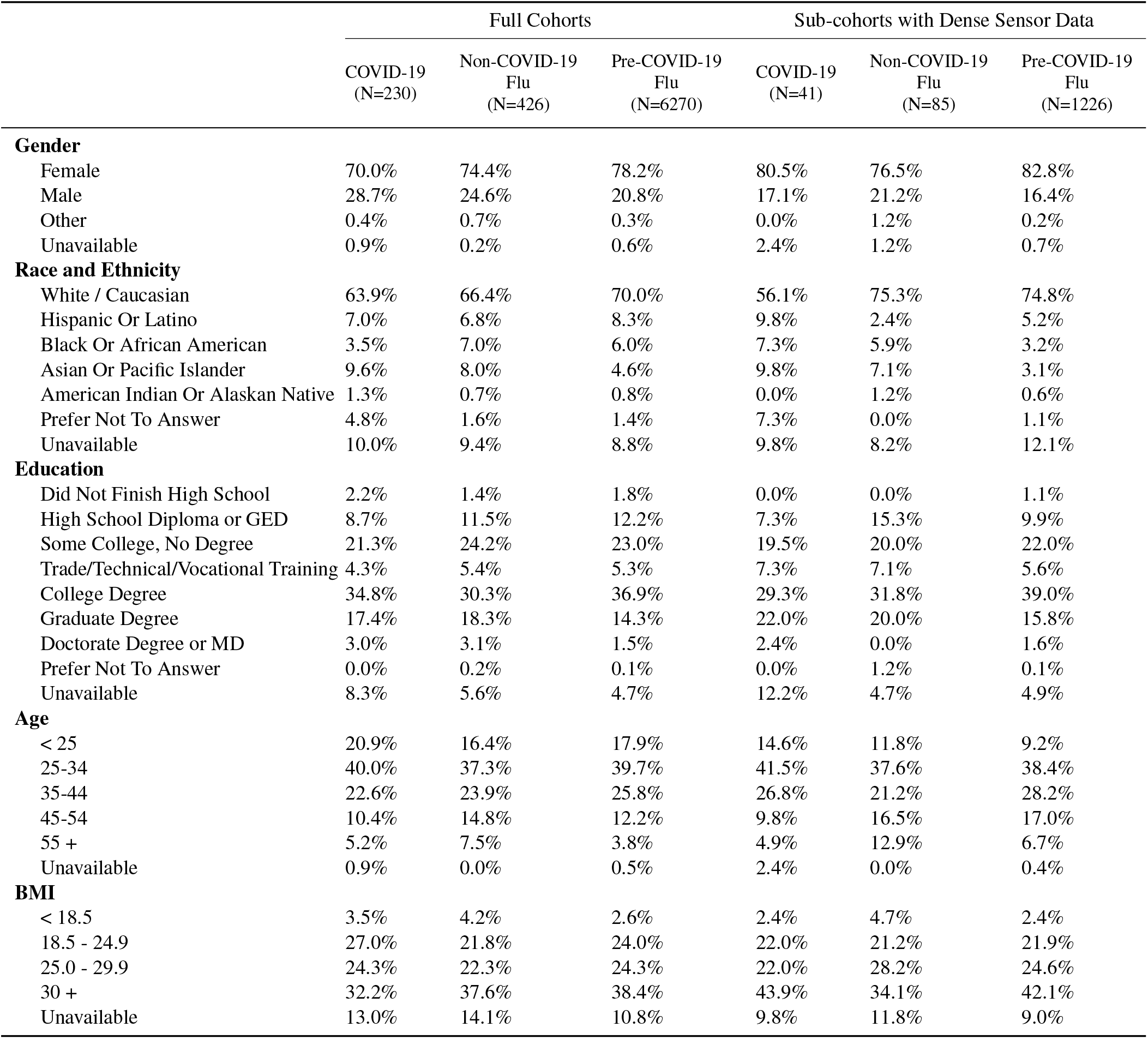
Demographic summaries for the full COVID-19, Non-COVID-19 Flu and Pre-COVID-19 Flu cohorts, as well as the subset of each cohort with dense steps, RHR, and/or sleep data.

|  | Full Cohorts |  |  | Sub-cohorts with Dense Sensor Data |  |  |
| --- | --- | --- | --- | --- | --- | --- |
|  | COVID-19<br>(N=230) | Non-COVID-19<br>Flu<br>(N=426) | Pre-COVID-19<br>Flu<br>(N=6270) | COVID-19<br>(N=41) | Non-COVID-19<br>Flu<br>(N=85) | Pre-COVID-19<br>Flu<br>(N=1226) |
| **Gender** |  |  |  |  |  |  |
| Female | 70.0% | 74.4% | 78.2% | 80.5% | 76.5% | 82.8% |
| Male | 28.7% | 24.6% | 20.8% | 17.1% | 21.2% | 16.4% |
| Other | 0.4% | 0.7% | 0.3% | 0.0% | 1.2% | 0.2% |
| Unavailable | 0.9% | 0.2% | 0.6% | 2.4% | 1.2% | 0.7% |
| **Race and Ethnicity** |  |  |  |  |  |  |
| White / Caucasian | 63.9% | 66.4% | 70.0% | 56.1% | 75.3% | 74.8% |
| Hispanic Or Latino | 7.0% | 6.8% | 8.3% | 9.8% | 2.4% | 5.2% |
| Black Or African American | 3.5% | 7.0% | 6.0% | 7.3% | 5.9% | 3.2% |
| Asian Or Pacific Islander | 9.6% | 8.0% | 4.6% | 9.8% | 7.1% | 3.1% |
| American Indian Or Alaskan Native | 1.3% | 0.7% | 0.8% | 0.0% | 1.2% | 0.6% |
| Prefer Not To Answer | 4.8% | 1.6% | 1.4% | 7.3% | 0.0% | 1.1% |
| Unavailable | 10.0% | 9.4% | 8.8% | 9.8% | 8.2% | 12.1% |
| **Education** |  |  |  |  |  |  |
| Did Not Finish High School | 2.2% | 1.4% | 1.8% | 0.0% | 0.0% | 1.1% |
| High School Diploma or GED | 8.7% | 11.5% | 12.2% | 7.3% | 15.3% | 9.9% |
| Some College, No Degree | 21.3% | 24.2% | 23.0% | 19.5% | 20.0% | 22.0% |
| Trade/Technical/Vocational Training | 4.3% | 5.4% | 5.3% | 7.3% | 7.1% | 5.6% |
| College Degree | 34.8% | 30.3% | 36.9% | 29.3% | 31.8% | 39.0% |
| Graduate Degree | 17.4% | 18.3% | 14.3% | 22.0% | 20.0% | 15.8% |
| Doctorate Degree or MD | 3.0% | 3.1% | 1.5% | 2.4% | 0.0% | 1.6% |
| Prefer Not To Answer | 0.0% | 0.2% | 0.1% | 0.0% | 1.2% | 0.1% |
| Unavailable | 8.3% | 5.6% | 4.7% | 12.2% | 4.7% | 4.9% |
| **Age** |  |  |  |  |  |  |
| < 25 | 20.9% | 16.4% | 17.9% | 14.6% | 11.8% | 9.2% |
| 25-34 | 40.0% | 37.3% | 39.7% | 41.5% | 37.6% | 38.4% |
| 35-44 | 22.6% | 23.9% | 25.8% | 26.8% | 21.2% | 28.2% |
| 45-54 | 10.4% | 14.8% | 12.2% | 9.8% | 16.5% | 17.0% |
| 55 + | 5.2% | 7.5% | 3.8% | 4.9% | 12.9% | 6.7% |
| Unavailable | 0.9% | 0.0% | 0.5% | 2.4% | 0.0% | 0.4% |
| **BMI** |  |  |  |  |  |  |
| < 18.5 | 3.5% | 4.2% | 2.6% | 2.4% | 4.7% | 2.4% |
| 18.5 - 24.9 | 27.0% | 21.8% | 24.0% | 22.0% | 21.2% | 21.9% |
| 25.0 - 29.9 | 24.3% | 22.3% | 24.3% | 22.0% | 28.2% | 24.6% |
| 30 + | 32.2% | 37.6% | 38.4% | 43.9% | 34.1% | 42.1% |
| Unavailable | 13.0% | 14.1% | 10.8% | 9.8% | 11.8% | 9.0% |

### Care-seeking Behavior

Although all patients had to report seeking medical care and being diagnosed with either Flu or COVID-19 to be included in the analyses, locations of medical care, hospitalization rates, and medication prescription rates differed significantly across the three cohorts (chi-squared tests of independence with a Bonferroni correction, all p<.001), and are summarized in Table 2. Compared to Non-COVID-19 Flu and Pre-COVID-19 Flu patients, COVID-19 patients were less likely to seek care at a primary care clinic (37.4% vs. 50.2% for Non-COVID-19 Flu, p =.002, and vs. 45.7% for Pre-COVID-19 Flu, p<.001) or urgent care facility (16.1% vs. 23.5% for Non-COVID-19 Flu, p =.026, and vs. 39.1% for Pre-COVID-19 Flu, p<.001) and more likely to seek care in an emergency room (17.0% vs. 8.2% for Non-COVID-19 Flu, p<.001, and vs. 6.9% for Pre-COVID-19 Flu, p<.001) or other location (37.4% vs. 50.2% for Non-COVID-19 Flu, p=0.002, and vs. 45.7% for Pre-COVID-19 Flu, p<.001). An informal review of the freeform text responses provided for the “other” location suggest that COVID-19 patients were more likely to seek care via telehealth services.

**Table 2.**
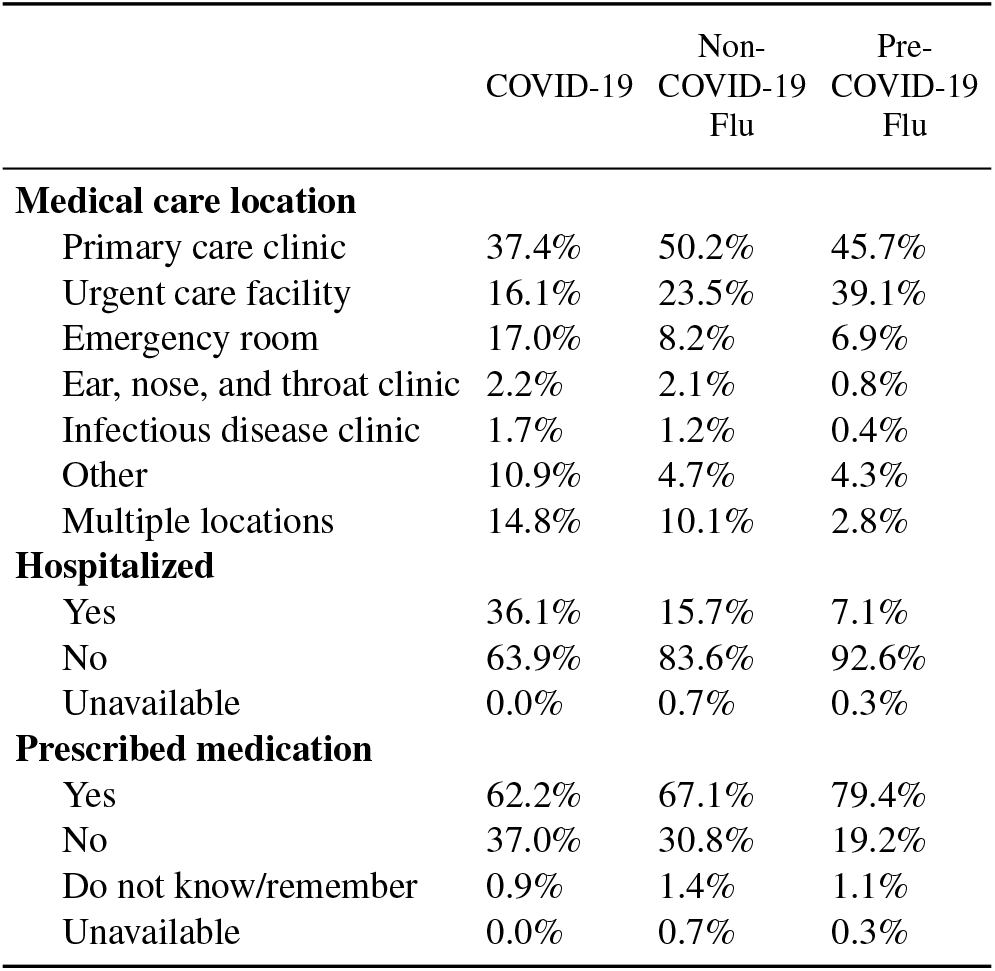
Summaries of medical care-seeking behaviors and outcomes for the COVID-19 (N=230), Non-COVID-19 Flu (N=426) and Pre-COVID-19 Flu (N=6270) groups, including where medical care was sought, whether patients were hospitalized, and whether they were prescribed medication.

COVID-19 patients were more likely to be hospitalized (36.1%) than Non-COVID-19 Flu (15.7%, p<.001) and Pre-COVID-19 Flu (7.1%, p< 0.001) patients. Interestingly, a greater proportion of patients with recent flu events were hospitalized than those with flu events earlier in the season (Non-COVID-19 Flu vs. Pre-COVID-19 Flu, p<.001).

Both the COVID-19 and Non-COVID-19 Flu cohorts were less likely to be prescribed medication than the Pre-COVID-19 Flu cohort (p<.001 and p<.001), but the medication rates between COVID-19 and Non-COVID-19 Flu patients did not differ significantly (p =.202).

### Symptom Prevalence

In addition to differences in medical care-seeking and outcomes, we also observed differences in self-reported symptoms between COVID-19 and the two cohorts of flu patients. A summary of symptom prevalence across cohorts is reported in Table 3. The most common symptoms across all ILI groups included cough, headache, body muscle ache, fatigue, and fever. The prevalence of symptoms was significantly different across the three cohorts (chi-squared test of independence, p<.001), confirming emerging literature and anecdotal reports (4, 10–13). All follow-up pairwise symptom comparisons were tested with two-proportion z-tests and a Bonferroni correction was applied for performing 33 tests.

**Table 3.** Summary of self-reported symptoms for the COVID-19 (N=230), Non-COVID-19 Flu (N=426) and Pre-COVID-19 Flu (N=6270) cohorts. Symptom prevalence refers to the percentage of the cohort reporting the symptom at any time during the ILI event and symptoms are sorted by most (top) to least (bottom) prevalence in the COVID-19 cohort. The day of peak symptom occurrence relative to illness onset corresponds to the maximum of a centered three-day rolling mean of day-by-day symptom prevalence for each cohort. Some symptoms (i.e., shortness of breath, chest pain/pressure, and anosmia) were only included in the updated survey and therefore are not available for the Pre-COVID-19 Flu cohort.

|  | Symptom Prevalence |  |  | Peak Symptom Day Relative to Illness Onset |  |  |
| --- | --- | --- | --- | --- | --- | --- |
|  | COVID-19 | Non-COVID-19 Flu | Pre-COVID-19 Flu | COVID-19 | Non-COVID-19 Flu | Pre-COVID-19 Flu |
| Cough | 84.3% | 71.6% | 85.1% | 5 | 3 | 3 |
| Headache | 71.3% | 68.1% | 74.3% | 4 | 3 | 3 |
| Body Muscle Ache | 66.1% | 67.1% | 80.8% | 4 | 2 | 2 |
| Shortness of Breath | 65.7% | 24.2% | NA | 6 | 8 | NA |
| Fatigue | 61.7% | 54.7% | 70.9% | 5 | 3 | 3 |
| Fever | 61.3% | 62.0% | 74.6% | 5 | 2 | 2 |
| Chills or Shivering | 53.5% | 55.4% | 69.3% | 4 | 3 | 2 |
| Sore Throat | 51.7% | 48.8% | 61.1% | 3 | 2 | 3 |
| Nasal Congestion | 49.6% | 49.3% | 65.4% | 5 | 2 | 3 |
| Chest Pain/Pressure | 49.6% | 19.7% | NA | 6 | 2 | NA |
| Sweats | 42.2% | 47.4% | 56.3% | 5 | 2 | 2 |
| Anosmia | 38.3% | 15.5% | NA | 7 | 3 | NA |
| Sneezing | 37.0% | 36.9% | 48.2% | 4 | 2 | 3 |

Compared to the Non-COVID-19 Flu cohort, patients with COVID-19 were significantly more likely to report experiencing cough (84.3% vs. 71.6%, two-proportion z-test, p<.001), loss of sense of smell (anosmia) (38.3% vs. 15.5%, p<.001), persistent pain or pressure in the chest (49.6% vs. 19.7%, p<.001), and shortness of breath or difficulty breathing (65.7% vs. 24.2%, p<.001). These symptoms are characteristic of COVID-19 and have been reported to differentiate COVID-19 from seasonal flu, although it’s important to note that, with the exception of cough, these symptoms are not necessarily common COVID-19 symptoms. For example, in the COVID-19 cohort, reports of chest pain were as common as reports of nasal congestion and sore throat, and reports of anosmia were as common as sneezing.

Compared to the Pre-COVID-19 Flu cohort, the COVID-19 cohort was significantly less likely to report experiencing body muscle ache, fever or feeling feverish, nasal congestion or runny nose, sneezing, chills or shivering, and sweats (all p<.001). The prevalence of several symptoms (i.e., shortness of breath, anosmia, and chest pain) could not be compared between the COVID-19 and Pre-COVID-19 Flu cohorts because they were not included in the original survey. Together, these findings are in line with recent work suggesting that the presentation of COVID-19 differs from other ILIs (2, 23), in particular with regard to anosmia, shortness of breath, coughing, fatigue, and muscle aches.

With the exception of headache, all symptoms were significantly less prevalent in the Non-COVID-19 Flu cohort relative to the Pre-COVID-19 Flu cohort. One possible reason for the difference in symptom presentations in the two flu cohorts is that the 2019–2020 flu season consisted of two waves of different flu strains: strain B (Victoria lineage) appeared earlier on and was followed by strain A (H1NI-pdm09) (24). According to the CDC, vaccines for the 2019–2020 season were well-matched against circulating strain A but not as well-matched against strain B (25), which could account for the more mild symptom presentation in the more recent flu cases in the Non-COVID-19 Flu cohort compared to the earlier cases in the Pre-COVID-19 Flu cohort.

In addition to the prevalence of individual symptoms across cohorts, we examined the prevalence of co-occurring sets of symptoms for the COVID-19 and Non-COVID-19 Flu cohorts (Figure 2). The Pre-COVID-19 Flu cohort was excluded from this analysis since only a subset of symptoms were available for this cohort. To reduce the complexity of the possible symptom sets, we included only the 5 most prevalent symptoms in each cohort, which resulted in a reduced set of 7 symptoms: cough, headache, fever, fatigue, body muscle ache, chills or shivering, and shortness of breath. The two most common symptom sets consisted of all symptoms, which was predominated by COVID-19 cases, and all symptoms except for shortness of breath, which was predominated by Non-COVID-19 Flu cases. The other symptom sets that were more indicative of COVID-19 than Non-COVID-19 Flu included the symptom pair of shortness of breath and cough, and all symptoms other than chills or shivering.

**Fig. 2.**
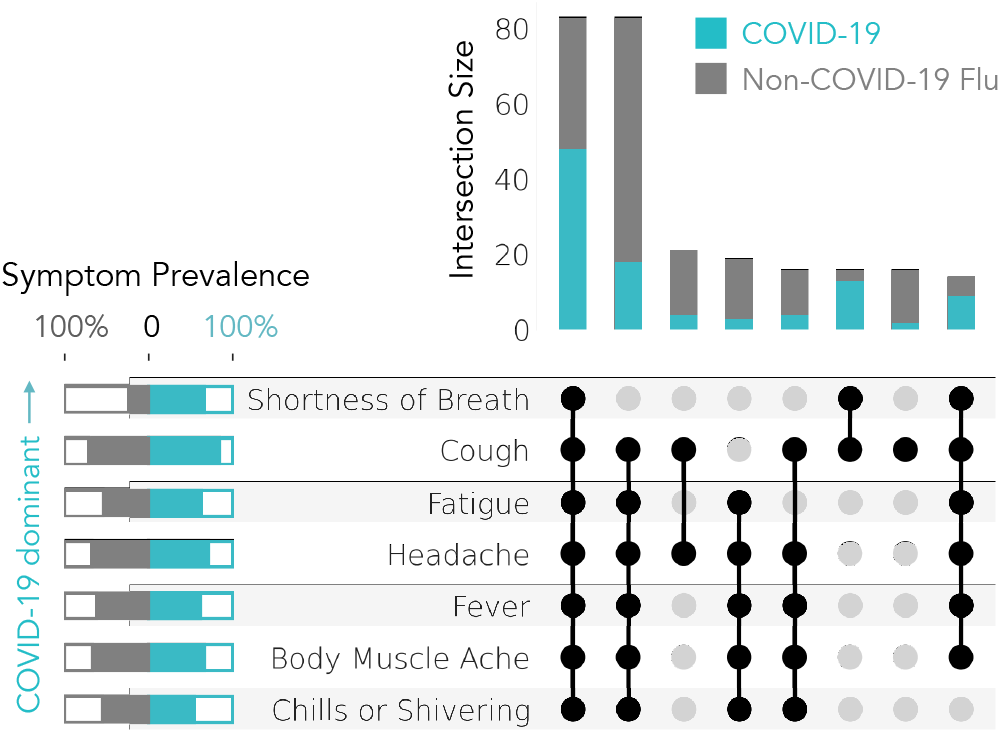
Co-occurrence of self-reported symptoms in COVID-19 cases (N=230; blue) or Non-COVID-19 Flu cases (N=426; gray). Only the top 5 most prevalent symptoms in each cohort are included in the symptom sets and only symptom sets that represent 2% or more of total COVID-19 and Non-COVID-19 Flu cases are plotted. Symptoms are sorted by their relative prevalence in COVID-19 (top) vs. Non-COVID-19 Flu (bottom) cases.

### Symptom Time Course

Patients reported the dates of illness onset and illness recovery, which were used to compute the duration of each ILI event in days (Figure 3). Duration of illness for COVID-19 cases tended to be longer than flu cases; COVID-19 illnesses lasted a median of 12 days, compared to 9 days for the Non-COVID-19 Flu cases and 7 days for the Pre-COVID-19 Flu cases.

**Fig. 3.**
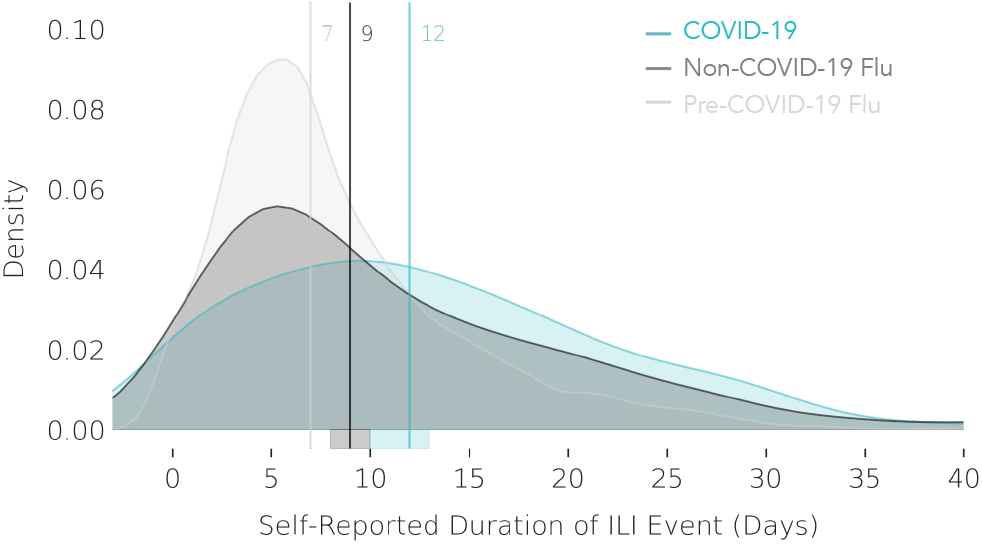
Self-reported illness duration in days for COVID-19 (N=230; blue), Non-COVID-19 Flu (N=426; gray), and Pre-COVID-19 Flu (N=6270, light gray) cases. Vertical lines denote the median illness duration.

Symptom prevalence across each cohort for each day after illness onset is illustrated in Figure 4 and the days of peak symptom occurrence for each cohort are reported in Table 3. In general, day-by-day symptom prevalence peaks later for the COVID-19 cases compared to the two groups of flu cases. With the exception of shortness of breath for Non-COVID-19 Flu cohort, all symptoms peak 2–3 days after illness onset in both flu cohorts. In contrast, COVID-19 symptoms peak 3–7 days after illness onset, with most symptoms peaking 4–5 days after illness onset. Some of the latest peaking symptoms are those that are most tightly associated with COVID-19, including shortness of breath, chest pain or pressure, and anosmia. Several symptoms present with prolonged durations in COVID-19, including fatigue, confirming anecdotal evidence (26).

**Fig. 4.**
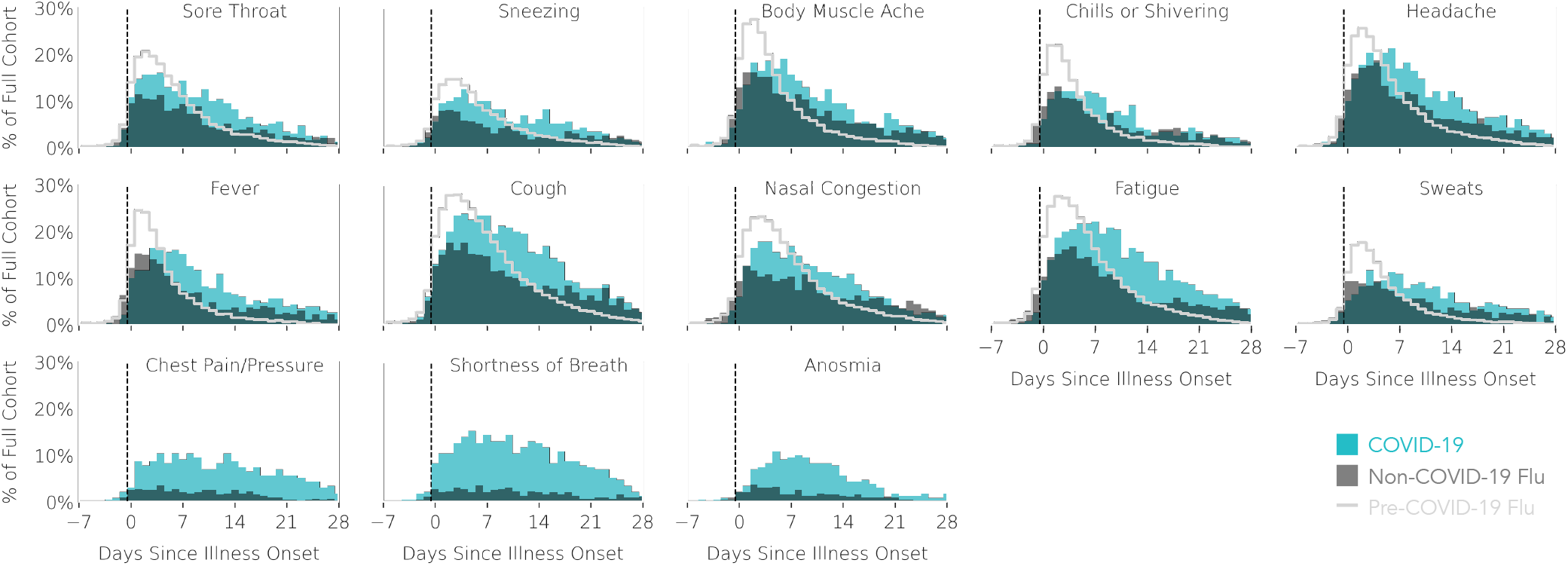
Self-reported symptom prevalence over time relative to illness onset (Day 0). Prevalence is reported as a percentage of the full cohort of COVID-19 cases (N=230; blue), Non-COVID-19 Flu cases (N=426; dark gray), or Pre-COVID-19 Flu cases (N=6270; light gray trace). Each subplot contains data for one symptom and symptoms are sorted by peak symptom occurrence (earliest to latest) for the COVID-19 cases.

### Wearable Sensor PGHD

Objective PGHD from commercial Fitbit sensors were available for at least one day between 2019–11–01 and 2020–05–13 for approximately 31% of all participants. A smaller subset of these participants met the criteria for sensor data density and were included in the analysis, including 41 (18%) COVID-19 patients, 85 (20%) Non-COVID-19 Flu patients, and 1,226 (20%) Pre-COVID-19 Flu patients. The demographics of the cohorts with dense sensor data are described in Table 1. The same two-step statistical testing procedure was applied to test for demographic differences among participants with (N=1,352) and without (N=5,574) dense sensor data. Compared to participants without dense Fitbit data, those with dense Fitbit data were more likely to be female (p< 0.001), white (p< 0.001), obese (30+ BMI, p=0.003), and in an older age group (45–54, p< 0.001; 55+, p< 0.001).

### Elevated RHR

Given the association between elevated RHR and the inflammatory immune system response (27, 28), we examined the prevalence of elevated RHR among the COVID-19, Non-COVID-19 Flu, and Pre-COVID-19 Flu around the ILI events (Figure 5. Elevated RHRs were defined as RHRs that surpassed participants’ mean RHR levels by 0, 0.5, or 1 standard deviation. In both the COVID-19 and Pre-COVID-19 Flu cohorts, we observe a spike in the prevalence of elevated RHR in the first few days after illness onset. We ran two-proportion z-tests to test 1) whether a greater proportion of COVID-19 patients had elevated RHRs around illness onset compared to before illness onset and 2) whether the prevalence of elevated RHR around illness onset differed between COVID-19 and Flu cases. The percent of COVID-19 patients with elevated RHRs (defined as a RHR >1 standard deviation above their personal mean RHR) was higher around the onset of COVID-19 (from Days –2 to 2) compared to the baseline period 5–10 days before illness onset (25% vs. 13%, p=0.005). The percent of the COVID-19 cohort with elevated RHR around illness onset was higher than that of the Non-COVID-19 cohort (16%, p=0.026), but did not differ from that of the Pre-COVID-19 cohort (22%, p=0.454).

**Fig. 5.**
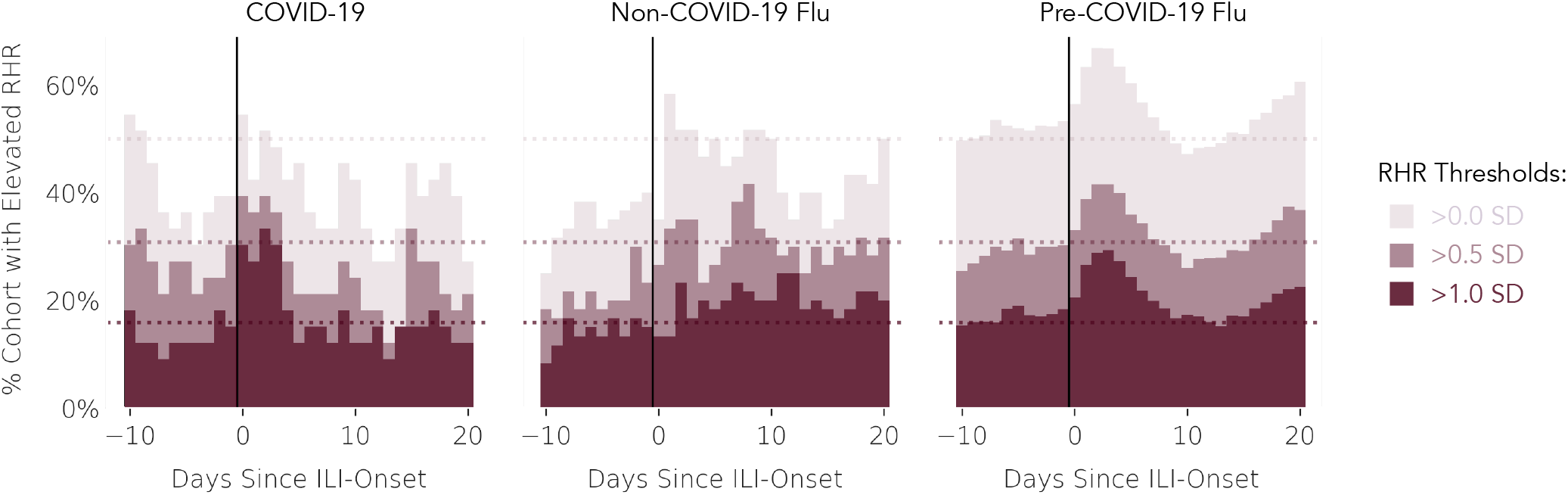
Fraction of participants with elevated RHR relative to illness onset (Day 0). Elevated RHR is defined as being greater than 1, 0.5, or 0 standard deviations (SD) of all daily RHRs available for that participant. Dotted horizontal lines denote the expected proportion of participants with an elevated RHR at each given threshold. RHR values were imputed and normalized (z-scored) before applying the thresholds. Cohorts refer to the subcohorts with dense RHR data: COVID-19 (N=33; left), Non-COVID-19 Flu (N=60; middle) and Pre-COVID-19 Flu (N=1025; right).

### ILI Impact Estimation

Symptoms reports analyzed in previous sections can be understood as an assessment of the deviation from how the participant normally feels. In this section we present an analysis of wearable data to derive an analogous of the concept of deviations from the norm during the ILI event that symptoms reports capture. The objective impact of ILI (COVID-19 or flu) was quantified as the excess observed in daily steps lost, additional minutes of sleep, and increased RHR as compared to the expected measurements recorded in the counter-factual scenario where symptoms were not present for that day and individual, as estimated from a model fit only to symptom-free days. We present aggregate time series of these excesses for each of the COVID-19, Non-COVID-19, and Pre-COVID-19 cohorts in Figure 6.

**Fig. 6.**
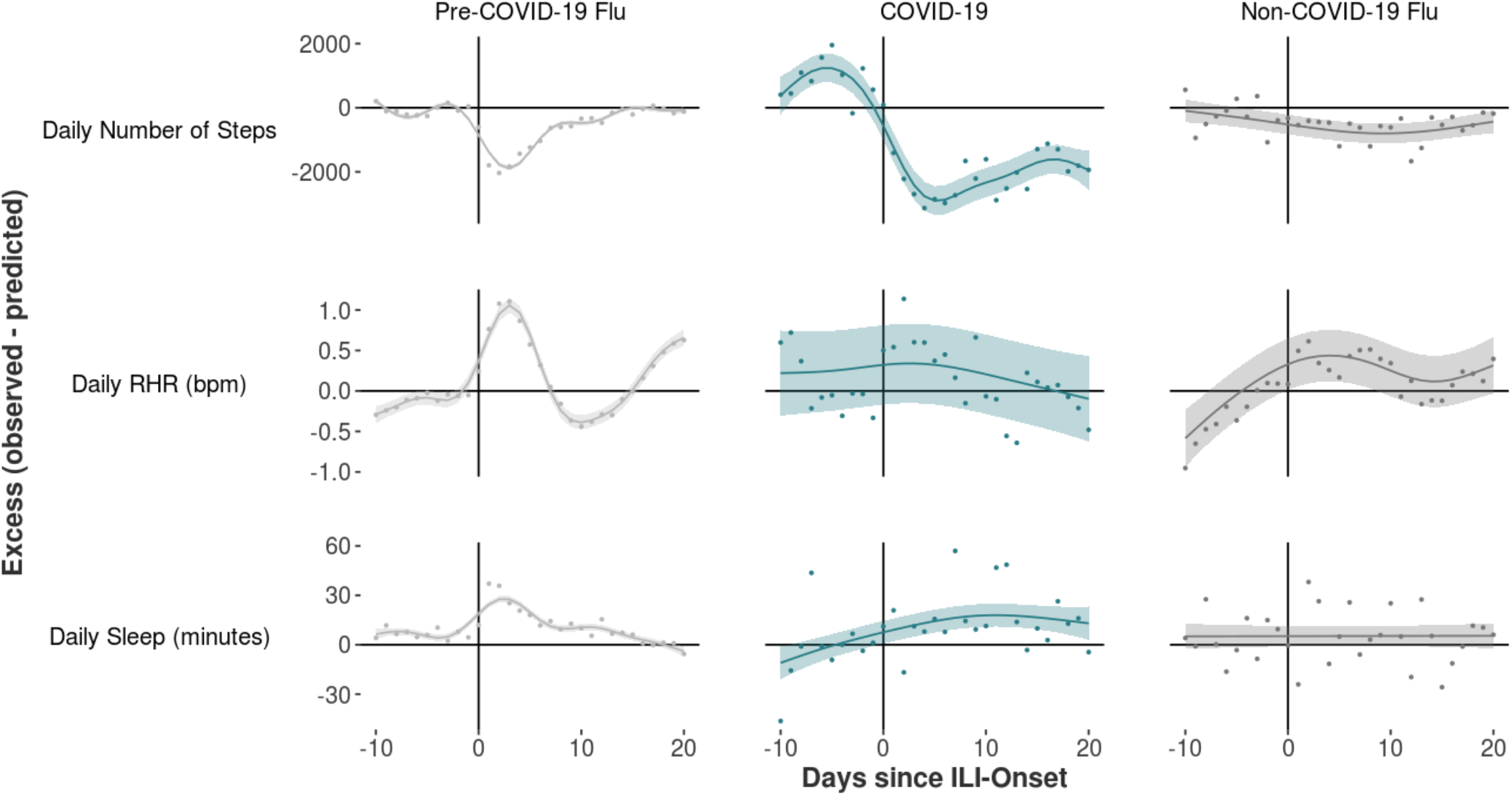
Deviations from typical healthy behavior and physiology observed during ILI events. Three measurement channels were studied: daily number of steps, daily mean RHR, and daily sleep minutes. Deviation from the norm was quantified as difference (excess) between observed values and estimates from a model fit only to symptom-free days (i.e., days outside the window of –10 through +20 days surrounding ILI-onset). Greater excess indicate greater deviations from typical behavior and physiology. In the steps analysis (top) Pre-COVID-19 N=1193, COVID-19 N=36, Non-COVID-19 N=80; in the RHR analysis (middle) Pre-COVID-19 N=1025, COVID-19 N=33, Non-COVID-19 N=60; and in the sleep analysis (bottom) Pre-COVID-19 N=979, COVID-19 N=35, Non-COVID-19 N=64.

Results are shown in Figure 6. Reductions in daily step counts are more marked and prolonged for COVID-19 as compares to Non-COVID-19 Flu and Pre-COVID-19 Flu. This is consistent with our observation of more extended symptoms and illness durations in the COVID-19 patients, but may also be explained by the adoption of more stringent self-imposed quarantine measures after COVID-19 diagnosis in addition to shelter-in-place. RHR has similar profiles for flu and COVID-19, suggesting the possibility to be used for real-time detection and monitoring of the time-course of the illness, but not necessarily forecasting before (self-reported) symptom onset or distinction of different ILIs. Sleep changes appear inconclusive, as the post-onset total sleep time increase observed for pre-stay-at-home flu may be explained by changes in sleeping schedule during sick days that would be less prominent when working from home.

## Conclusions

We present the first read-out of PGHD including longitudinal symptoms reports and linked data from commercial wearables for 6,926 diagnosed flu and 230 diagnosed COVID-19 patients remotely collected in real-life settings.

We describe symptoms across different ILIs, in order to build a better understanding of COVID-19 presentation and contextualize it with flu. Specific symptoms, including chest pain, shortness of breath or anosmia, as well as combinations of these symptoms (e.g., shortness of breath and coughing) were more prevalent in COVID-19 as compared to Non-COVID-19 Flu. Other symptoms, including fatigue and cough, were more pronounced later after illness onset. Generally, patients reported longer COVID-19 illnesses (median of 12 days) than Non-COVID-19 and Pre-COVID-19 Flu illnesses (9 and 7 days, respectively).

Our results show that COVID-19 patients are far more likely to seek emergency care, and are more likely to be hospitalized. This observation supports the severity with which COVID-19 infection is taken, and the scaling up of the medical response to the pandemic. We also observe however that COVID-19 patients are less likely to be prescribed medication, indicating the current dearth of available proven treatments, and further underlining the need for remote monitoring and containment (29).

Differences in self-reported symptoms are supported by data from the subset of our cohort who also contributed wearable sensors PGHD. We observed that activity (measured as lost steps) was reduced longer and more prominently for COVID-19 patients as compared to other groups. This is consistent with the observed longer illness durations for COVID-19 generally, but could also reflect orders to self-quarantine. Consistent with this line of reasoning, 89% of the COVID-19 cohort reported being told by a medical provider to self-quarantine compared to only 57% of the Non-COVID-19 Flu cohort.

We observe a significantly increased fraction of participants with elevated RHR measurements in the 2 days surrounding ILI symptoms onset. This has previously been observed for other ILIs (7), and is also observed for COVID-19 patients. If validated in large and representative populations (30), our findings suggest the potential for PGHD to support remote monitoring of infectious disease patients, with opportunities ranging from improve resource allocation for further remote diagnostic, to inform population-level early-warning systems based on geo-localized aggregate symptoms.

The prominence of passively collected sensor data among other PGHD data sources requires further investigation. The key observation from our findings is that although sensor data can be a general, real-time trigger for ILI recognition and tracking, subjective symptom information is required to provide specificity in distinguishing COVID-19 from flu and likely other ILI. While disease-specificity may be improved by additional data sources such as pulse oximetry, respiration rate, electrodermal activity, but may still ultimately require further interactions with the individual. This underlines that although passive monitoring may be a key pillar of a largescale monitoring of ILIs generally, a deeper understanding of the cost/benefits of the interactions that may be triggered, and, in general of the decisions driven by it, is needed in order to assess any clinical utility.

### Biases & Limitations

The studied cohorts come from convenience samples that are not representative of the US population at large, in particular we note that African-Americans, alongside older individuals, are underrepresented in our cohort therefore our findings may not generalize. Increasing access and usage of these tools in these risk groups (31) is therefore of critical importance. Differences in the rate of the occurrence across demographic groups and disease severity levels have not been investigated, though preliminary findings point to possible differences with other ILIs (23, 32). Largescale connected populations offer the ability to reduce this divide and examine the impact COVID-19 is having across demographic and geographic groups, helping to highlight vulnerable populations and target care delivery (4).

It is also clear that even our approach may underestimate severity, due to participants not reporting symptoms or not wearing sensors in days when symptoms are most severe, or during hospitalization events (see Figure S3). The conclusions presented here may therefore even be conservative.

Additionally, we recognize that our analyses do not immediately translate to real-time implementation, due to lag in data collection that comes from sensor synchronization and data synchronization. Lags in our dataflow are nevertheless small compared to the gains in symptom detection and reporting compared to canonical practice.

Clearly the COVID-19 pandemic is still in its infancy, and although our work on surveying annual waves of ILI infections is well established, testing is currently reserved for patients with severe symptoms, or strongly suspected to be infected with COVID-19. This could create a self-fulfilling prophecy where patients that match the current assumptions about presentation are more likely to be tested, and therefore more likely to confirm current thinking in our symptom reporting. For example, a patient with non-canonical COVID-19 symptoms may not get tested but simply be told they have another ILI. This further underlines the criticality of widespread testing to develop a canonical presentation.

We are also aware that in our selection of ILI events, for each participant we select COVID-19 events or the most recent ILI event. This biases our analysis towards later calendar dates when sensor data is most affected by social distancing. For this reason we have included a chronologically parallel group of Non-COVID-19 Flu patients. A second issue is that we could be missing participants’ most severe ILI events, which could have happened earlier in the season. We will continue to monitor symptomatic and behavioral changes associated with COVID-19 and non-COVID-19 ILIs as more events are captured and as guidance on social distancing and stay-at-home measures are relaxed. Further analysis will focus on how strongly these measures confound our observations.

### Outlook

These subjective (self-reported) and objective (via commercial wearables) PGHD allow us to learn about symptom presentation, care-seeking behavior, and contextualize COVID-19 as compared to flu. As more data is being collected, further work will focus on increasing the breadth of participation, and examining other PGHD signals which may support early detection and differentiation of ILIs. To accelerate learnings and findings generalizability it would be desirable that the many ongoing study initiatives could participate in the creation of a data consortium to facilitate access to cross-study harmonized datasets to a broader audience of qualified researchers, following the example of data consortia that are being created for Real-World Data (33).

As care becomes more decentralized and telehealth becomes more prominent (34), PGHD can become a valuable tool on an individual level as patients transition in and out of care (35). Equally, taken in aggregate, PGHD can provide insights into public health, such as hotspot detection and real time-monitoring for public health interventions, crucial in monitoring effectiveness of reopening and enabling contact tracing (29, 36).

The vast majority of learnings about COVID-19 have come from real world data sources such as EHR, claims, etc. PGHD can be an crucial addition to the real world data arsenal, adding a large-scale understanding of early signals, several days before impact is seen at centers of care. As the COVID-19 pandemic continues to develop, and as future annual ILI waves arrive, understanding and correctly reacting to symptom presentation will be critically important. These results support not only an emerging picture that COVID-19 has a distinct presentation, but highlight the power of PGHD, digital health, and connected populations in broadly and remotely monitoring health status.

## Data Availability

De-identified study data will be made available to qualified researchers on the Sage Synapse platform in September 2020.

## ACKNOWLEDGEMENTS

The authors would like to thank Raghu Kainkaryam for technical input, and Christine Lemke, Malay Gandhi, Stephanie Jones for feedback on the manuscript. The authors would also like to thank Dr. Stephen Friend and the team at 4YouandMe for the invaluable partnership and discussions.

## Supplementary Note 1: Institutional Review Board

This study received expedited review and IRB approval from Solutions IRB (Protocol ID #2018/11/8). Waiver of informed consent was granted by the IRB. Prior to each questionnaire, participants were notified about how their survey responses and behavioral data will be used for research purposes through a disclosure.

## Supplementary Note 2: Questionnaire

### Weekly 1-Click Item

1. Have you experienced flu-like symptoms in the past 7 days (such as fever, chills, cough, shortness of breath, and/or headache)? If you had flu-like symptoms in the past 7 days, but have recovered, please still answer YES.

a. Yes [Symptom Experience Survey]
b. No [Infection Risk Factors Survey]

### Symptom Experience Survey

1. What is your current zip code? (Where you live and spend the majority of your time) *If you are currently staying in a different location for an extended period of time, please enter your current zip code*.

a. numeric 5-digit entry
2. When did you first begin experiencing flu-like symptoms? If you don’t recall the exact date, please provide the best estimate.

a. calendar date selection
3. As of today, do you feel that you have completely recovered from your illness?

a. Yes
b. No [IF Q3=A, THEN Q4] [IF Q3=B, THEN THEN SKIP TO Q5]
4. When did you feel you were completely recovered from your illness? If you don’t recall the exact date, please provide the best estimate.

a. calendar date selection
5. We’d like to know more about the symptoms you experienced. Looking back over the past 7 days, did you have any of the following symptoms? Please select all that apply.

a. Cough
b. Body/Muscle Ache
c. Fever or feeling feverish
d. Chills or shivering
e. Sweats
f. Headache
g. Sore throat or itchy/scratchy throat
h. Feeling more tired than usual
i. Nasal congestion or runny nose
j. Sneezing
k. I did not experience any flu-like symptoms
l. Other [IF Q4=K, SURVEY END]
6. We’d like to know more about the symptoms you experienced. Looking back over the past 7 days, please indicate on which days you felt the following symptoms.

a. Matrix carry forward symptoms; check all that apply: Today, Yesterday, 2 days ago, 3 days ago, 4 days ago, 5 days ago, 6 days ago
7. Looking back over the past 7 days, did you have any of the additional symptoms below

a. Shortness of breath and/or difficulty breathing
b. Persistent pain or pressure in the chest
c. Loss of sense of smell
d. None of the above [IF Q7=A-C, THEN Q8] [IF Q7=D, THEN THEN SKIP TO Q9]
8. Looking back over the past 7 days, please indicate on which days you felt the following additional symptoms.

a. Matrix carry forward other symptoms; check all that apply: Today, Yesterday, 2 days ago, 3 days ago, 4 days ago, 5 days ago, 6 days ago
9. Thinking about your flu-like symptoms over the last 7 days, on what day did you feel the worst?

a. Today
b. Yesterday
c. 2 days ago
d. 3 days ago
e. 4 days ago
f. 5 days ago
g. 6 days ago
10. Did you seek medical attention from a healthcare provider at a clinic or urgent care facility for this flu or flu-like illness?

a. Yes
b. No [IF Q10=A, THEN Q11] [IF Q10=B, THEN SKIP TO Q23]
11. Where did you seek care from a healthcare provider?

a. Primary care clinic (e.g. family medicine, internal medicine)
b. Urgent care facility
c. Emergency room (ER)
d. Ear, nose, and throat (otolaryngology) clinic
e. Infectious disease clinic
f. Other
12. Did the healthcare provider diagnose you as having the flu?

a. Yes
b. No
c. I don’t know / I can’t remember
13. Did the healthcare provider perform any of the following tests? Select all that apply.

a. Nasal swab
b. Throat swab
c. Symptoms only (no lab test)
d. I don’t know / I can’t remember
e. Other (please specify)
14. Did the healthcare provider diagnose you as having coronavirus disease (also known as COVID-19)?

a. Yes
b. No
c. I am waiting for my diagnosis
d. I don’t know / I can’t remember
15. Did you take any of the following tests for your coronavirus diagnosis? Select all that apply.

a. Nasal swab
b. Throat swab
c. Blood test
d. Spit test / kit
e. Symptoms only (no lab test)
f. I don’t know / I can’t remember
g. Other (please specify)
16. Where did you take the COVID-19 diagnostic test?

a. In a clinic or hospital
b. At a drive through testing facility
c. At home testing kit
d. Other (please specify)
e. None of the above
17. Were you hospitalized as a consequence of this flu or flu-like illness? *Hospitalization is when you leave the emergency room (ER) and are admitted to the inpatient hospital based on a doctor’s order. Even if you stayed overnight in the ER, this is not considered a hospitalization*.

a. Yes
b. No
18. Were you told to self-quarantine (stay in your home without leaving for any reason) by a medical professional?

a. Yes
b. No
c. I don’t know / I can’t remember
19. Did a healthcare provider prescribe any medications to treat or manage your current symptoms?

a. Yes
b. No
c. I don’t know / I can’t remember [IF Q19=A, THEN Q20] [IF Q19=B, THEN SKIP TO Q23]
20. Which of the following medications were you prescribed to treat or manage your symptoms? Select all that apply.

a. Xofluza (baloxavir marboxil)
b. Tamiflu (oseltamivir)
c. Relenza (zanamivir)
d. Antibiotics (Z-pak, amoxicillin, Augmentin, doxycycline)
e. Other
21. When did you take your first dose of [CARRY FORWARD MEDICATION NAME]? Please enter the date in MM/DD/YYYY format.

a. Date entry
22. Did you ever miss any doses or decide not to take [CARRY FORWARD MEDICATION NAME]? a.

a. I missed at least one dose of this medication
b. I did not take any doses of this medication
c. I did not miss any doses of medication
d. I don’t know / I can’t remember
23. Did you take any over-the-counter (non-prescription) medications to treat or manage your current symptoms in the past 7 days?

a. Yes
b. No
c. I don’t know / I can’t remember [IF Q23=A, THEN Q24] [IF Q23=B, THEN SKIP TO Q25]
24. Which of the following over-the-counter (non-prescription) medications did you personally decide to take to treat or manage your current symptoms in the past 24 hours? Select all that apply.

a. Fever reducers or pain relievers (ibuprofen, aspirin, Advil, Tylenol, Aleve, acetaminophen)
b. Cough suppressants (Delsym, Robitussin, dextromethorphan)
c. Chest or mucus decongestants (Mucinex, guaifenesin)
d. Nasal decongestants (Sudafed, Sudafed PE, Afrin, Flonase, phenylephrine, pseudoephedrine, fluticasone propionate)
e. I don’t know / can’t remember
f. Other
25. How many people (other than yourself) live in your household?

a. 0
b. 1
c. 2
d. 3
e. 4
f. 5
g. 6
h. 7
i. 8
j. 9
k. 10
l. > 10
26. Have any members of your household (other than yourself) experienced flu-like illness this flu season?

a. Yes
b. No
c. I live alone [IF Q26=A, THEN Q27] [IF Q26=B or C, THEN SKIP TO Q29]
27. How many members of your household, by age group listed below, have experienced flu-like symptoms during this flu season (September 2019 to today)? If no household member in your household experienced symptoms within an age group please enter 0. [numeric entry]

a. Number of household members 0–4 years old experiencing flu-like symptoms
b. Number of household members 5–17 years old experiencing flu-like symptoms
c. Number of household members 18–49 years old experiencing flu-like symptoms
d. Number of household members 50–64 years old experiencing flu-like symptoms
e. Number of household members 65+ years old experiencing flu-like symptoms
28. Have any members of your household been diagnosed with coronavirus disease (also known as COVID-19?

a. Yes
b. No
29. Have you been in close contact with anyone outside your household (e.g., family members, friends, coworkers, acquaintances) who has experienced flu-like symptoms recently? *Close contact can include direct physical contact, face-to-face contact for longer than 15 minutes, exchange of bodily fluids, or being within 6 feet of the person for more than 15 minutes*.

a. Yes, within the last 7 days
b. Yes, within the last 14 days
c. Yes, over 14 days ago
d. No
e. I don’t know / I’m not sure
30. Have you recently been in contact with someone who was diagnosed with coronavirus? *Close contact can include direct physical contact, face-to-face contact for longer than 15 minutes, exchange of bodily fluids, or being within 6 feet of the person for more than 15 minutes*.

a. Yes, within the last 7 days
b. Yes, within the last 14 days
c. Yes, over 14 days ago
d. No
e. I don’t know / I’m not sure
31. Did you miss school or work due to your illness?

a. No, I did not miss any school or work during my illness
b. I missed 1 day of school or work
c. I missed 2 days of school or work
d. I missed 3 days of school or work
e. I missed more than 3 days of school or work
f. Illness occurred on a weekend or other day
g. off
h. I am retired and/or school or work days don’t apply to me
i. I don’t know / I don’t remember
32. Looking back over the past 7 days, which days have you practiced social distancing or isolation behaviors (e.g., working remotely, limited the time spent in crowds, increasing the amount of time spent at home)? Please select all that apply.

a. Today
b. Yesterday
c. Two days ago
d. Three days ago
e. Four days ago
f. Five days ago
g. Six days ago
h. I did not practice social distancing in the last 7 days
33. Did you receive the flu vaccine (sometimes called the flu shot) this flu season (September 2019 to today)?

a. Yes
b. No
c. I don’t know / I can’t remember
34. Did you receive the flu vaccine last flu season? (September 2018 – March 2019)

a. Yes
b. No
c. I don’t know / I can’t remember
35. Please select the statement below that describes whether you typically get a flu shot (or another form of flu vaccine).

a. I never have gotten a flu shot
b. I rarely get a flu shot
c. I get a flu shot every year
d. I sometimes get a flu shot
36. Have you recently traveled on an airplane?

a. Yes, within the last 7 days
b. Yes, within the last 14 days
c. Yes, over 14 days ago
d. No
37. Have you recently participated in any large public gatherings of over 250 people (e.g., concerts, sporting events, amusement parks)?

a. Yes, within the last 7 days
b. Yes, within the last 14 days
c. Yes, over 14 days ago
d. No
38. Are you or one of your household members a healthcare worker (i.e., doctor, dentist, nurse, nurse’s aid, paramedic, physician’s assistant, home healthcare aid, hospital worker, pharmacist, or other type of healthcare worker)? Please select all that apply.

a. I am, and I am currently working
b. I am, but I am NOT currently working
c. One of my household members is, and they are currently working
d. One of my household members is, but they are NOT currently working
e. No one in my household is a healthcare worker

### Infection Risks Factors Survey

1. What is your current zip code? (Where you live and spend the majority of your time) *If you are currently staying in a different location for an extended period of time, please enter your current zip code*.

a. numeric 5-digit entry
2. Have you recently traveled on an airplane?

a. Yes, within the last 7 days
b. Yes, within the last 14 days
c. Yes, over 14 days ago
d. No
3. Have you recently participated in any large public gatherings of over 250 people (e.g., concerts, sporting events, amusement parks)?

a. Yes, within the last 7 days
b. Yes, within the last 14 days
c. Yes, over 14 days ago
d. No
4. Are you or one of your household members a healthcare worker (i.e., doctor, dentist, nurse, nurse’s aid, paramedic, physician’s assistant, home healthcare aid, hospital worker, pharmacist, or other type of healthcare worker)? Please select all that apply.

a. I am, and I am currently working
b. I am, but I am NOT currently working
c. One of my household members is, and they are currently working
d. One of my household members is, but they are NOT currently working
e. No one in my household is a healthcare worker
5. Have any members of your household (other than yourself) experienced flu-like illness this flu season?

a. Yes
b. No
c. I live alone [IF Q5=A, THEN Q6] [IF Q5=B or C, THEN SKIP TO Q8]
6. How many members of your household, by age group listed below, have experienced flu-like symptoms during this flu season (September 2019 to today)? If no household member in your household experienced symptoms within an age group please enter 0. [numeric entry]

a. Number of household members 0–4 years old experiencing flu-like symptoms
b. Number of household members 5–17 years old experiencing flu-like symptoms
c. Number of household members 18–49 years old experiencing flu-like symptoms
d. Number of household members 50–64 years old experiencing flu-like symptoms
e. Number of household members 65+ years old experiencing flu-like symptoms
7. Have any members of your household been diagnosed with coronavirus disease (also known as COVID-19?

a. Yes
b. No
8. Have you been in close contact with anyone outside your household (e.g., family members, friends, coworkers, acquaintances) who has experienced flu-like symptoms recently? *Close contact can include direct physical contact, face-to-face contact for longer than 15 minutes, exchange of bodily fluids, or being within 6 feet of the person for more than 15 minutes*.

a. Yes, within the last 7 days
b. Yes, within the last 14 days
c. Yes, over 14 days ago
d. No
e. I don’t know / I’m not sure
9. Have you recently been in contact with someone who was diagnosed with coronavirus? *Close contact can include direct physical contact, face-to-face contact for longer than 15 minutes, exchange of bodily fluids, or being within 6 feet of the person for more than 15 minutes*.

a. Yes, within the last 7 days
b. Yes, within the last 14 days
c. Yes, over 14 days ago
d. No
e. I don’t know / I’m not sure
10. Looking back over the past 7 days, which days have you practiced social distancing or isolation behaviors (e.g., working remotely, limited the time spent in crowds, increasing the amount of time spent at home)? Please select all that apply.

a. Today
b. Yesterday
c. Two days ago
d. Three days ago
e. Four days ago
f. Five days ago
g. Six days ago
h. I did not practice social distancing in the last 7 days

## Supplementary Note 3: Methods

Figure S1 describes the filtering process for the selection of the analysis dataset from the input dataset, including Survey Filtering and Inference and Reconciliation of distinct ILI events.

**Fig. S1.**
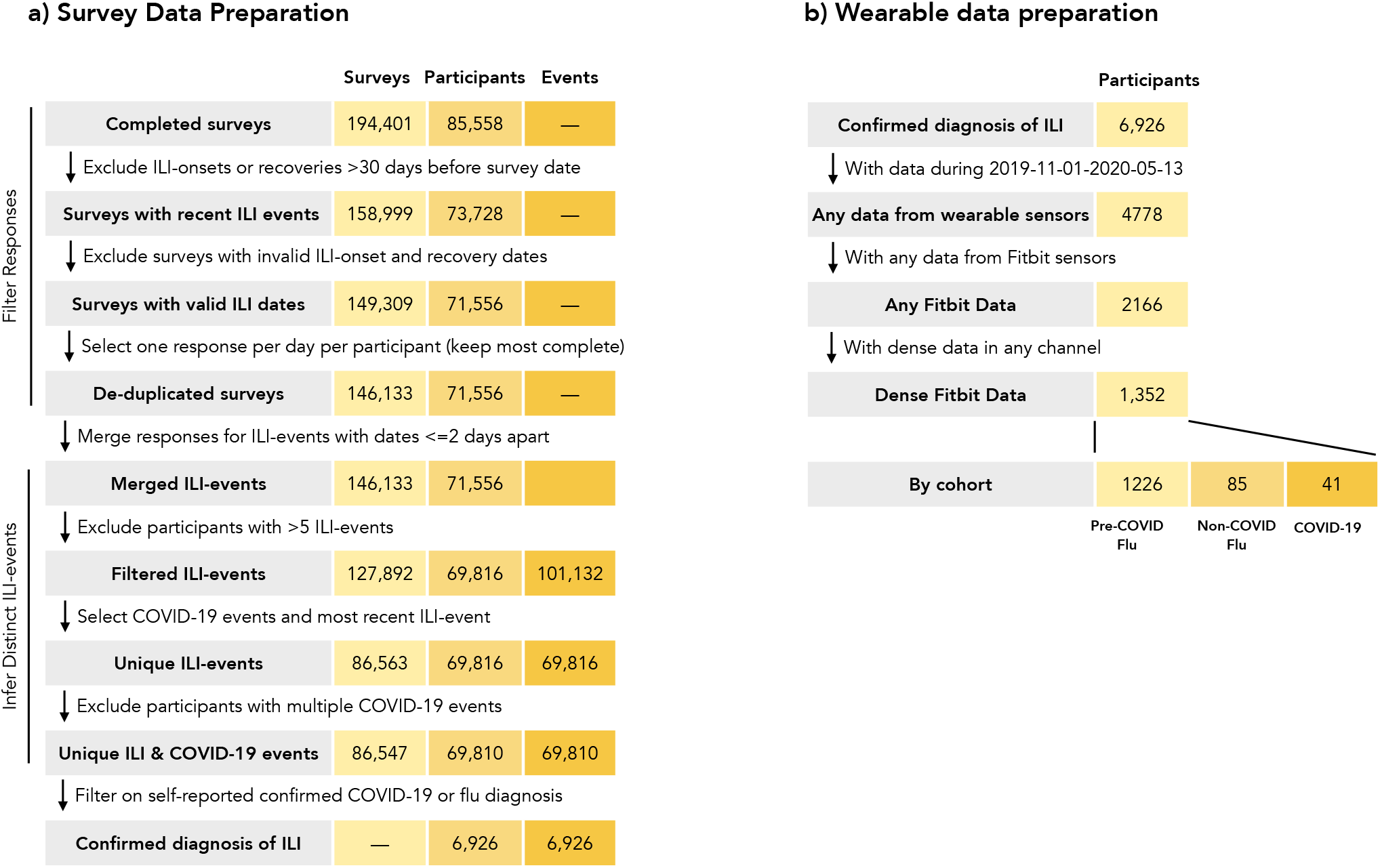
(a) Flow diagram for preparing survey data for analysis. Data preparation consisted of filtering survey responses and merging responses that correspond to the same ILI event. (b) Flow diagram for preparing Fitbit wearable data. Participants with data too sparse were filtered out.

### Survey Filtering

Survey responses with self-reported illness onset dates or recovery dates that occurred 30 or more days before the survey completion date were excluded, leaving 158,999 survey responses from 73,728 unique participants. Survey responses with invalid illness onset and/or recovery dates (defined as dates occurring after the survey date or responses in which the illness recovery date occurred before the illness onset date) were also removed, leaving 149,309 survey responses from 71,556 unique individuals. Finally, the set of survey responses was restricted to one survey per participant per day. If one participant attempted more than one survey in a given day, the less complete survey was excluded.

### Inference of Distinct ILI Events

Participants could submit new survey responses as frequently as once per week, with no maximum limit. Therefore, individual symptom trajectories for an ILI event had to be inferred by concatenating and reconciling multiple surveys responses, for example, if the participant were midway through their illness when they submitted their first survey their next survey could describe the second half of their illness.

We inferred ILI events by merging multiple surveys from the same participant with date ranges encompassing symptoms onset and recovery that overlapped or were separated by no more than 2 days. Participants with more than 5 ILI events were removed, eliminating 16,878 surveys and 1,639 participants, and leaving 126,014 survey responses, corresponding to 99,604 distinct ILI events and 69,034 participants.

This gives the set of discrete ILI events per participant, from which we will select only 1 for analysis. If a participant has a diagnosed COVID-19 ILI event, that event is selected, otherwise the most recent ILI event is selected. This process removes a further 40,357 surveys and 30,567 distinct ILI events, and 0 participants. Participants reporting multiple non-overlapping diagnosed COVID-19 events were then excluded (excluding 7 survey responses, 6 distinct ILI events, and 3 participants).

### Reconciliation of Merged Survey Responses

At this point, we have one ILI event per participant, corresponding to 85,650 surveys for 69,031 distinct ILI events across 69,031 participants. We then reconcile responses to derive a single value per item. For example, the date of onset and recovery are taken as the earliest and latest reported date for that ILI event, respectively.

Flu events drawing from multiple surveys responses may have differing symptoms reports for the same calendar date. Such day-level values (e.g., symptoms reported for a specific day) were collapsed if identical, and if not, the survey submitted on the date closest to the calendar date was used. Participants were also allowed to report annotations, for example “the worst day”, during a given event. These are highly subjective, thus all responses were retained, with a given date coded as “*one of* the worst days” if the participant indicated as such in any survey.

For event-level categorical features, the algorithm described in Figure S1 was used to collapse surveys to a single response. Numerical event-level features, for example the number of household members who have experienced ILI symptoms, were aggregated by taking the maximum value reported. All other features which could not be reconciled were simply aggregated as concatenated unique values.

## Supplementary Note 4: Comorbidity Prevalence

Table S1 describes self-reported comorbidities observed in our ILI cohorts.

**Table S1.** Prevalence of self-reported co-morbidities for the COVID-19 (N=230), Non-COVID-19 Flu (N=426), and Pre-COVID-19 Flu (N=6270) cohorts.

|  | COVID-19 | Non-COVID-19<br>Flu | Pre-COVID-19<br>Flu |
| --- | --- | --- | --- |
| Anxiety | 65 (28.3%) | 122 (28.6%) | 1915 (30.5%) |
| Depression | 62 (27.0%) | 104 (24.4%) | 1868 (29.8%) |
| Asthma | 56 (24.3%) | 79 (18.5%) | 1247 (19.9%) |
| Migraines | 50 (21.7%) | 75 (17.6%) | 1225 (19.5%) |
| Chronic Pain | 30 (13.0%) | 40 (9.4%) | 572 (9.1%) |
| Hypertension | 17 (7.4%) | 46 (10.8%) | 718 (11.5%) |
| PCOS | 16 (7.0%) | 16 (3.8%) | 315 (5.0%) |
| GERD | 15 (6.5%) | 46 (10.8%) | 599 (9.6%) |
| Mental Health (Excluding Depression/Anxiety) | 15 (6.5%) | 32 (7.5%) | 499 (8.0%) |
| Insomnia | 15 (6.5%) | 37 (8.7%) | 541 (8.6%) |
| Sleep Apnea | 11 (4.8%) | 20 (4.7%) | 335 (5.3%) |
| Restless Leg Syndrome | 10 (4.3%) | 12 (2.8%) | 231 (3.7%) |
| Type 2 Diabetes | 10 (4.3%) | 10 (2.3%) | 254 (4.1%) |
| Hypo- or Hyperthyroidism | 9 (3.9%) | 32 (7.5%) | 418 (6.7%) |
| Fibromyalgia | 9 (3.9%) | 16 (3.8%) | 215 (3.4%) |
| High Cholesterol | 9 (3.9%) | 21 (4.9%) | 352 (5.6%) |
| Gestational Diabetes | 8 (3.5%) | 13 (3.1%) | 187 (3.0%) |
| Cancer | 6 (2.6%) | 13 (3.1%) | 165 (2.6%) |
| Arrhythmia | 6 (2.6%) | 5 (1.2%) | 165 (2.6%) |
| Psoriasis | 5 (2.2%) | 14 (3.3%) | 147 (2.3%) |
| Type 1 Diabetes | 4 (1.7%) | 6 (1.4%) | 63 (1.0%) |
| Rheumatoid Arthritis | 4 (1.7%) | 7 (1.6%) | 135 (2.2%) |
| Stroke | 3 (1.3%) | 3 (0.7%) | 31 (0.5%) |
| Heart Attack | 2 (0.9%) | 3 (0.7%) | 28 (0.4%) |
| IBS or IBD | 2 (0.9%) | 9 (2.1%) | 94 (1.5%) |
| COPD | 2 (0.9%) | 4 (0.9%) | 61 (1.0%) |
| Seasonal Allergies | 2 (0.9%) | 5 (1.2%) | 109 (1.7%) |
| Lupus | 1 (0.4%) | 0 (0.0%) | 7 (0.1%) |
| Coronary Heart Disease | 1 (0.4%) | 0 (0.0%) | 13 (0.2%) |
| Multiple Sclerosis | 1 (0.4%) | 4 (0.9%) | 30 (0.5%) |
| Alzheimer's Disease | 1 (0.4%) | 1 (0.2%) | 3 (0.0%) |
| Heart Failure | 0 (0.0%) | 5 (1.2%) | 27 (0.4%) |
| Neurodegenerative | 0 (0.0%) | 1 (0.2%) | 4 (0.1%) |
| Arthritis | 0 (0.0%) | 3 (0.7%) | 37 (0.6%) |
| Osteoporosis | 0 (0.0%) | 6 (1.4%) | 51 (0.8%) |

## Supplementary Note 5: Symptom Labels

Table S2 describes the labels and associated descriptions used in our surveys for this work.

**Table S2.** Full symptom descriptions included in the survey for each abbreviated symptom label. The Chest Pain/Pressure, Shortness of Breath, and Anosmia symptoms were only included in the updated survey.

| Symptom Label | Symptom Description in Survey |
| --- | --- |
| Cough | Cough |
| Headache | Headache |
| Body Muscle Ache | Body/Muscle Ache |
| Fatigue | Feeling more tired than usual |
| Fever | Fever or feeling feverish |
| Chills or Shivering | Chills or shivering |
| Sore Throat | Sore throat or itchy/scratchy throat |
| Nasal Congestion | Nasal congestion or runny nose |
| Sweats | Sweats |
| Sneezing | Sneezing |
| Chest Pain/Pressure | Persistent pain or pressure in the chest |
| Shortness of Breath | Shortness of breath and/or difficulty breathing |
| Anosmia | Loss of sense of smell |

## Supplementary Note 6: Symptom Reporting

Figure S2 describes the percentage of each ILI cohort reporting daily symptoms between one week prior and 4 weeks post symptom onset. Figure S3 describes the percentage of observed symptom reporting for hospitalized and non-hospitalized COVID-19 cohorts, between one week prior and 4 weeks post symptom onset.

**Fig. S2.**
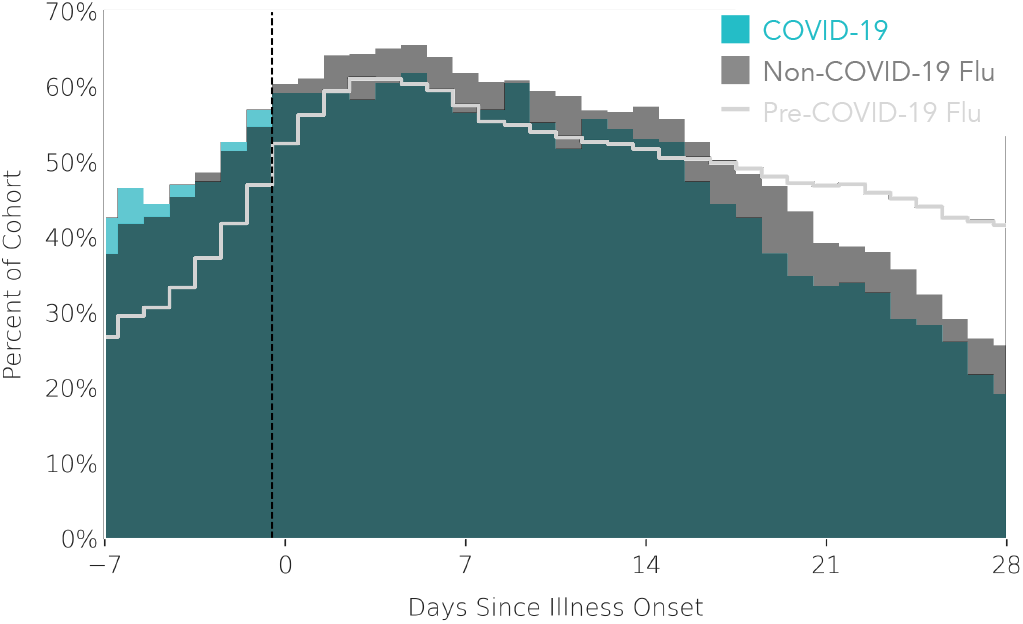
Percentage of COVID-19 (N=230; blue), Non-Covid Flu (N=426; gray), and Pre-Covid Flu (N=6270, light gray trace) cohorts with symptom reports for days –7 to 28 since illness onset.

**Fig. S3.**
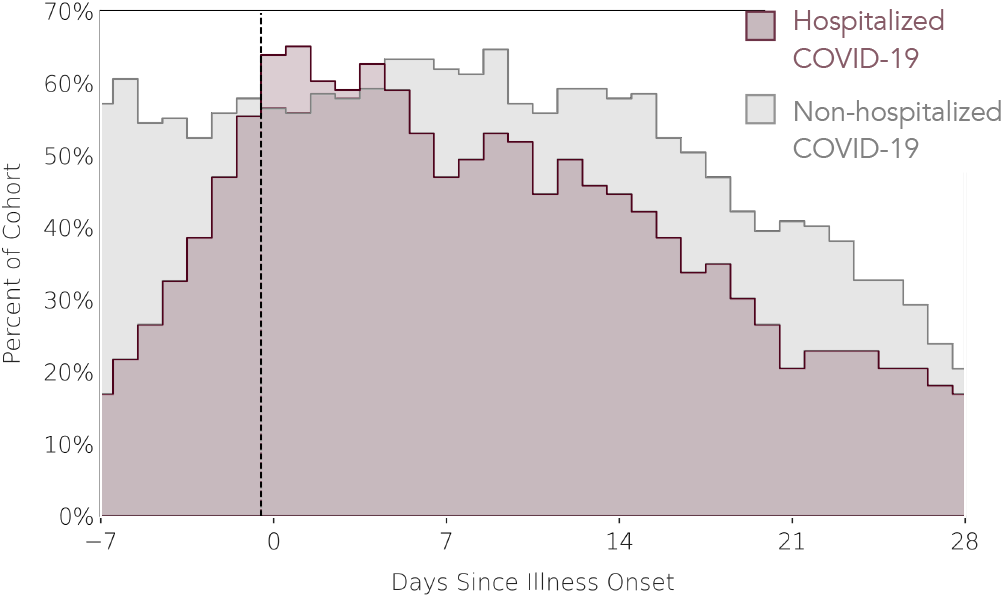
Percentage of the Hospitalized (N=83, purple) and Non-hospitalized (N=147, gray) COVID-19 sub-cohorts with symptom reports for days –7 to 28 since illness onset.

## Supplementary Note 7: Sensor Data Coverage

A summary of coverage of wearable sensor data over the course of the study is visualized in Figure S4.

**Fig. S4.**
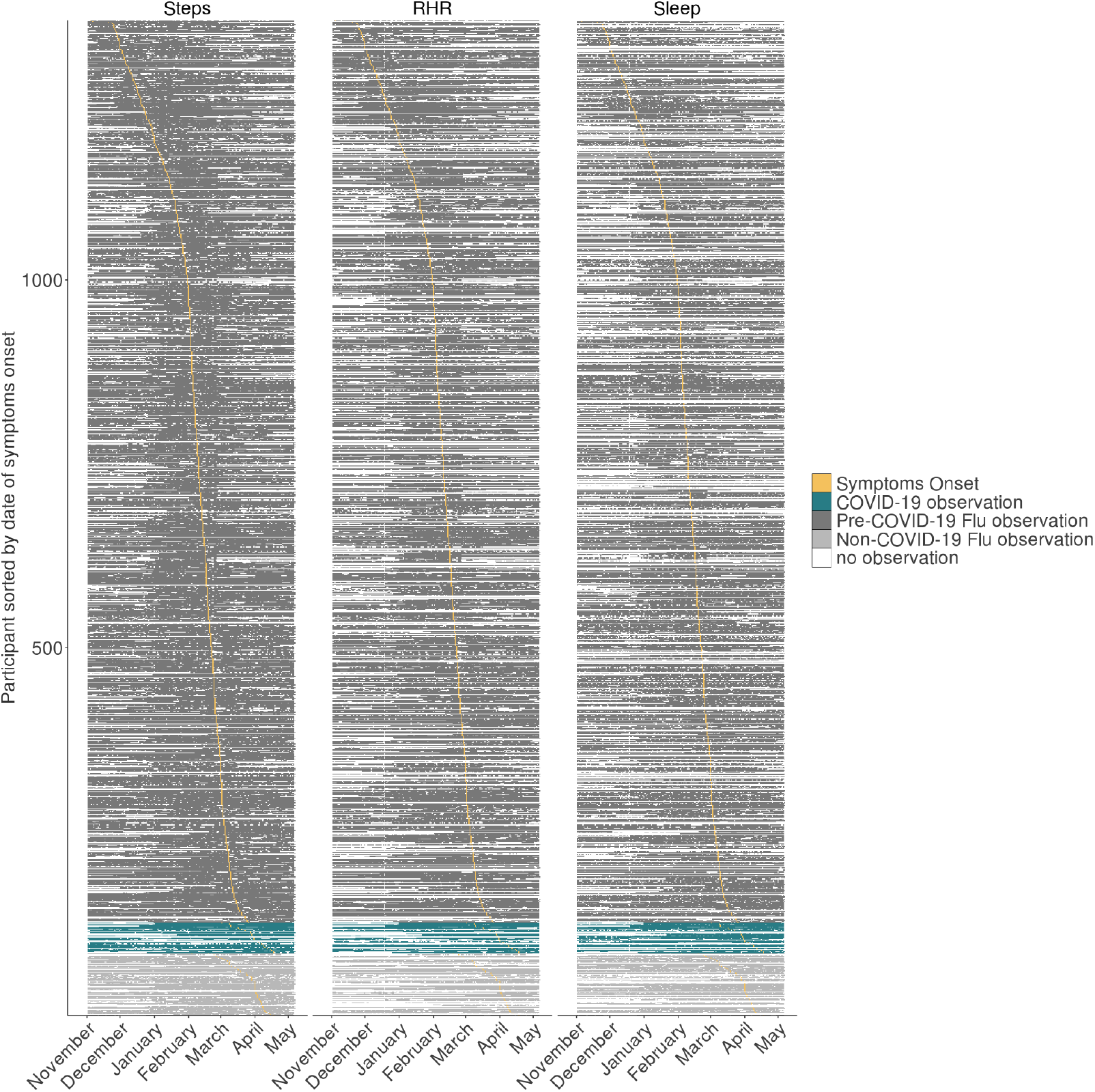
Coverage of Fitbit steps, sleep, and RHR data on each calendar date of the study, color-coded by cohort. Each row is one participant (ordered by date of ILI-onset) and each column is one calendar date. Shaded days indicate that wearable data was recorded on that day from that participant. Days highlighted in yellow indicate the ILI onset dates.

## Supplementary Note 8: Data Availability

De-identified study data will be made available to qualified researchers on the Sage Synapse platform (37) in September 2020.

## Footnotes

1 We conservatively allow 2 days for physiological changes before any symptom is reported, and up to 2 days after the onset of symptoms as the time horizon within which actions can be taken that would not be taken otherwise, without information from the wearable device)

2 Median time from exposure to development of symptoms is 5 days for COVID-19, (19)

